# “It’s hard to keep a distance when you’re with someone you really care about” — a qualitative study of adolescents’ pandemic-related health literacy and how Covid-19 affects their lives

**DOI:** 10.1101/2021.06.17.21257667

**Authors:** Kirsti Riiser, Kåre Rønn Richardsen, Kristin Haraldstad, Sølvi Helseth, Astrid Torbjørnsen

## Abstract

**Purpose:** The aim of this study was to explore how adolescents accessed, understood, appraised, and applied information on pandemic preventive measures, how their lives were impacted by long-lasting regulations and how they described their quality of life.

**Methods:** A qualitative design with focus group interviews was used to elaborate on the quantitative survey results obtained and analyzed in a previous survey study from the first phase of the Covid-19 pandemic. Five focus groups with seventeen adolescents were conducted digitally during the second pandemic phase in November and December 2020. The interview data were analyzed with directed content analysis.

**Results:** The adolescents reported using traditional media and official websites as sources for Covid-19 information. They engaged in preventive behavior, and washing hands and keeping a distance from strangers had become a habit. However, not being physically close to friends felt strange and unpleasant. The measure most frequently discussed was limiting social contact, which was a constant struggle. No one disputed the authorities’ guidelines and rules, but the social restrictions caused boredom and despair, particularly due to interrupted schooling and missed opportunities to engage in life events, and freely socialize with friends.

**Conclusion:** The adolescents gave an overall impression of being health literate, which corresponds well with the results from our previous survey study. Their descriptions of how they translated protective measures into their everyday lives demonstrate that they took responsibility and accepted personal costs for the collective good. However, life with social restrictions decreased their quality of life.

## Introduction

Effective prevention of Covid-19 infection during the pandemic relies on the public adhering to key preventive measures. To stop the virus from spreading, health authorities and governments around the world have implemented comprehensive measures to limit social contact. Adolescents are more likely to socialize in close peer groups, which makes it particularly important that they comply with the regulations. The application of guidelines and regulations requires individual decision-making and agency, which again partly relies on adolescents’ level of health literacy. Health literacy is commonly described as the ability to find, understand, appraise, and apply health information [1]. During the pandemic, the health authorities have consistently presented simple and practical information about basic protective measures like hand-washing and keeping a physical distance. However, the regulations on school closures, social events, gatherings, traveling and leisure-time activities have rapidly been shifting based on the infection situation and the evolving scientific knowledge about the virus. Translating fast-changing requirements into actions in one’s own life may be challenging. In our previous cross-sectional study of Norwegian youth carried out during lockdown in the initial phase of the pandemic, we found that the participating adolescents appeared health literate and were loyal to the health authorities’ guidelines [2]. Other cross-sectional studies from the same period showed that adolescents engaged in protective measures [3, 4], that higher disease knowledge was associated with higher risk perception and that those with higher risk perception were more likely to comply [5]. However, few studies so far have investigated health literacy among young people in the context of the pandemic in more depth.

Protective measures like distancing and school closure may particularly impact young people by causing isolation and loneliness which are shown to be associated with adverse mental health outcomes [6, 7]. Moreover, concern about possible symptoms or fear that relatives will become infected may cause stress and worry [8]. Our previous study found that, compared to norm values, the participating adolescents’ health-related quality of life (HRQoL) was low [2]. Several studies have been published describing similar findings, showing that shortly after the pandemic outbreak young people reported lower HRQoL and more mental health problems [7, 9-11]. Recent research also shows that the proportion of young people reporting low quality of life and poor mental health was higher later in the pandemic compared to the early phase [12]. However, limited qualitative research exists exploring how adolescents reflect on their own contributions to limiting the spread of the virus and how such radical and long-lasting measures impact their quality of life.

The aim of this study is to explore adolescents’ pandemic-related health literacy and investigate how protective measures affect their quality of life. By doing so, the present qualitative study elaborates on the findings of our cross-sectional study of adolescents’ health literacy and HRQoL during the initial phase of the Covid-19 pandemic [2]. We investigate how the situation is experienced by adolescents in the second phase of the pandemic and how their lives are affected by the pandemic over time. The research questions are as follows: How do adolescents access, understand, appraise, and apply information on pandemic preventive measures? How are their lives impacted by the long-lasting regulations and how do they describe their quality of life?

## Methods

### Study design

We used qualitative methods with focus group interviews to elaborate on the quantitative survey results obtained and analyzed during an earlier phase of the pandemic [2]. These two studies together constitute a sequential mixed-methods design as the qualitative study builds on the findings of the quantitative study. The qualitative data and analysis refine and explain the survey results by exploring adolescents’ views in more depth [13]. Collecting quantitative data and then interviewing adolescents about the same topics at a later point of time serves two purposes. First, the adolescents were given the opportunity to reflect on their own experiences when discussing the topics investigated quantitatively. Secondly, early experiences can be contrasted with later experiences and thus provide knowledge of how adolescents act and react within different phases of the pandemic. The study was conducted and reported in accordance with the Consolidated Criteria for Reporting Qualitative Research (COREQ).

### Recruitment and setting

Altogether, 5 focus groups with 3–4 participants were recruited from 7 higher secondary schools in south-eastern Norway. We aimed at a heterogeneous sample consisting of boys and girls and students attending different study programs. As there has been geographical variation in social restrictions, we recruited from rural areas with less spread of the virus, as well as Oslo, the capital of Norway. From the start of the pandemic until the time of data collection, Oslo had the highest relative number of reported cases throughout the pandemic. Recruitment was based on direct approach to principals at different schools and on snowballing. A weblink with information on the study was issued to the adolescents who volunteered to participate. After a consent form was signed digitally along with contact information, each participant was invited to attend one online focus group meeting.

### Data collection

The focus group interviews were conducted in November and December 2020. Pairs of researchers (KR and KRR or AT and KRR) led and moderated the interviews. KR (female) and KRR (male) are both physiotherapists and AT (female) is a registered nurse. The remaining two researchers, KH (female) and SH (female) are registered nurses and SH is also a public health nurse. All the researchers involved in the study are experienced researchers and hold PhDs. A semi-structured interview guide with open ended questions was developed containing the following themes: sources for information on Covid-19 (where to find, how to understand, how to assess, and how to apply), how they adhered to the regulations, how their everyday life was organized around the protective measures, and how they experienced life during a pandemic. The adolescents were asked to reflect on the situation right now and how things might have changed over the course of the pandemic. Initially, the participants were given information about the study, the reasons for doing the research, as well as a short presentation about the research group members and their interest in the research topic. Information on age, gender and whether the informant lived with cohabitants, in a city, town, or rural area was collected. No relationship was established before the commencement of the study, but the interviewers aimed at creating a relaxed atmosphere for the participants during the interviews. The interviews were performed via Zoom, and audio files were uploaded using an encrypted link to TSD, a service for sensitive data [14]. The interviews were transcribed verbatim in TSD using VLC Media Player. The transcripts were anonymity-checked by two researchers in the project group before exported for analysis outside TSD.

### Analysis

Qualitative content analysis was applied. This is defined as a research method for the subjective interpretation of the content of text data through the systematic classification process of coding and identifying themes or patterns [15]. When prior research exists about a phenomenon that would benefit from further description as was the case here, directed content analysis is a suitable approach. The previously conducted study [2] and the commonly applied understanding of health literacy and HRQoL provided directions for the analysis [15]. To answer the first research question, all text was read through and given a predetermined code representing one of the four components commonly defined as health literacy: finding, understanding, appraising and applying health information [1]. The latter (applying health information) was subsequently coded relating to the main protective measures of handwashing, distancing, and limiting social contacts. A new code for knowledge of protective measures was identified and added. The same procedure was followed to answer the second research question. We applied predetermined codes derived from HRQoL as operationalized by the Kidscreen-instrument which was used in our previous study: physical well-being, psychological well-being, autonomy and parent relations, peers and social support, and school environment [16]. After the initial coding process, meaning units were selected and sorted before the content was abstracted and thematized. Quotes from the participants were extracted either to illustrate common patterns or to display diversity in the material. All the authors read the material to obtain an overall understanding of the data, while the analysis was performed by the first and last authors. The results were discussed among all the authors. NVivo 12 software was used to classify, sort, and arrange the text in the process of analyzing the data.

### Ethics

The study was presented to the Norwegian Regional Committee for Medical and Health Research Ethics. The Committee concluded that the study is not covered by the Health Research Act because the project does not make use of or bring forth new personal health information or sensitive data. The study protocol was reviewed by the Data Protection Official for Research to ensure that the project was in accordance with the Personal Data Act and the Personal Health Data Filing System Act (reference number 210900). Informed written consent was obtained digitally from all the participants. The audio files were deleted after they were transcribed.

## Results

Altogether, 17 adolescents, 9 girls and 8 boys, from 7 different schools participated in 5 focus group interviews with 3–4 participants in each group (Table 1). The adolescents were in the age range 15– 19 years. Only one participant had been infected with the sars-CoV-2 virus. Except for group 3, the groups consisted of participants who knew each other well. The interviews lasted from 28 to 62 minutes.

**Table 1.** Characteristics of the study participants

| Focus group | Number of participants | Gender | Residency | Study program | Action level* |
| --- | --- | --- | --- | --- | --- |
| 1 | 4 | 4 girls | Town | Vocational | Yellow |
| 2 | 3 | 3 boys | Town | Vocational | Yellow |
| 3 | 3 | 3 girls | Village | General | Yellow |
| 4 | 4 | 2 boys, 2 girls | City | General | Red |
| 5 | 3 | 3 boys | City | General | Red |

\*Schools are graded by green, yellow and red action levels: green level = schools can be organized as normal; yellow level = a class is considered a cohort and measures are to be taken to limit the contacts between cohorts; red level = students are divided into small groups, which leads to restricted school attendance in order to keep the distance required [17].

In Norway, a national lockdown during spring 2020 followed by a relaxation of the regulations during the summer before the restrictions increased again during autumn and winter. By December 2020, well into what is described as the second wave, the health authority’s guidelines for hand hygiene, social distancing, and travel bans were supplemented with face masks being made mandatory in public places in the large cities. There were strict regulations on indoor leisure activities which had been closed for periods [17].

The adolescents in groups 1 and 2 attended a school that had been running on a yellow level the entire autumn, and they had not been experiencing online teaching since the lockdown. Group 3’s school had been on the red action level several times and had recently returned from the red to the yellow action level. The latter two groups included adolescents from various schools in Oslo, the capital of Norway, all of which had been on the red action level with mainly online teaching and less physical attendance since the beginning of November 2020.

The content analysis resulted in the following themes: “information everywhere,” “understanding and confusion,” “complying with the rules,” “missing social interactions,” and “loss of opportunities.” Each theme is described in the following sections.

### Information everywhere

The participants gave several examples of sources for pandemic-related information. The majority mentioned traditional media such as TV and newspapers, and several said they tried to watch the health authorities’ and governments’ broadcast press briefings. Adolescents in all the groups highlighted the Norwegian Institute of Public Health’s web pages as a preferred site and the most reliable source of coronavirus-related information. Several underlined the importance of checking the source and publication dates of online articles about the pandemic and subsequent measures. Some said that their parents usually provided updates on new regulations, while the adolescents who were actively involved in sports relied on their coaches to provide information on relevant restrictions and precautions. Several participants found that the health authorities’ short public health messages conveyed through social media like Snapchat and Twitter were effective for keeping updated. Schools stood out as significant providers of advice and information. All the groups referred to their school’s web page as the number one place to look for locally relevant information:

> *“In my school there are a lot of updates on what to do and how to handle things. I feel that the school is my most important source for corona information*.*” (Girl, group 4)*

Although the adolescents requested and searched for information on specific regulations when needed, they also expressed a feeling of information overload:

> “*I mean, it’s everywhere. No matter where you are, it’s: corona, corona!” (Girl, group 4)*

The latter statement is an illustration of what seemed to lie beneath the informants’ utterances: that information was primarily received rather than actively retrieved and that instructions and advice on protective measures was presented to them in all areas of life all the time.

### Understanding and confusion

The adolescents were well aware of their school’s action level and the general advice on hand washing, distancing and use of face masks. However, several respondents were confused about the rules pertaining to limiting social contact. They expressed uncertainty about the number of guests allowed into private homes, as the national and local guidelines sometimes differed and changed from week to week depending on the number of confirmed cases in the area. The lack of coherence in the application of regulations in school seemed to evoke annoyance. One girl described how a measure to reduce the possibility of close contact with fellow students did not work as intended:

> *“At school, we’re not allowed to sit on the benches in the lobby. They are covered with plastic. The school administration tries to limit the number of contacts by removing places to sit, which doesn’t work at all. I don’t understand the logic behind that rule. As long as we sit next to each other in the classrooms, we might as well be close in the lobby. It’s not as if the virus stops being contagious in the classroom*.*” (Girl, group 3)*

Another informant seemed frustrated by the different rules relating to various classes and locations at school:

> *“In the classroom, we have to stay one meter apart. But in drama class, we’re allowed to touch, hug and roll around on the floor together. So, it feels wasted to keep the distance in some classes while in others we don’t. It feels silly*.*” (Girl, group 4)*

Some of the adolescents experienced that there had been a shift from advice and recommendations to rules and regulations and that things seemed more organized during autumn 2020 than during spring 2020. As the number of cases increased after the summer, they found the health authorities’ measures more specific and imperative:

> *“Now it’s more like straightforward and directed messages, so I try my best just to stay home and skip everything and hope to catch up later*.*” (Boy, group 5)*

Although the participants were critical of some of the schools’ efforts to minimize the spread of the virus, and questioned their relevance, no one disputed the government’s general protective advice and regulations. They seemed to have accepted that strict regulations are necessary to reduce the transmission of the virus to protect the elderly and those at risk of serious disease.

### Complying with the rules

Among the measures most frequently mentioned were handwashing or using sanitizer, distancing, and limiting the number of social contacts. At the time of data collection, facemasks were mandatory only in public places in the largest cities and were therefore not a major topic in the interviews. Most of the adolescents said they were more afraid of the virus in the first phase of the pandemic than by the end of 2020, and that they used to pay more attention to hand hygiene reminders in the beginning. Several said that, although they still washed their hands more frequently than before, they were less concerned for their own sake. However, one girl said that washing hands had become an obsession while one of the boys described how touching surfaces in public places felt unpleasant.

All the adolescents reported that they hugged and shook hands less than usually and that they really missed it. Some said that, during the first lockdown, it was hard to remember to keep a distance, but now it had become a habit. Others said that although it is easier to keep a distance from strangers, for example on the bus, it still felt awkward and difficult not being physically close to friends:

> *“Of course, it’s harder to keep a distance when you are with someone you really care about. Simply because you love them. It’s like you’re attracted to your friends in a way. So, it is a real struggle. Not to mention saying “no” when someone takes the initiative to give me a hug*.*” (Girl, group 4)*

The measure most frequently mentioned and discussed was limiting social contact. This seemed like a constant fight for most of the adolescents, but they also gave examples of how they managed to adhere to the maximum number of individuals per group allowed indoors. One girl explained how the larger group split up based on gender or the old group structure from lower secondary school. One boy said that making plans with fewer people had become the new normal:

> *“You just know that there are no gatherings with lots of people, so lately it has almost become a routine not to hang out in larger groups*.*” (Boy, group 2)*

Several described the discomfort of being the one who exceeded the limit of participants allowed per group and thus had to stay home:

> *“At the beginning I felt like only some of us took the responsibility, so I had to think about it for everyone, like ok, we’re too many, so I can’t join, but all the others are going, so it was kind of… everyone else was joining, but I couldn’t because then we were too many*.*” (Girl, group 1)*

The adolescents spoke about how they felt punished for following the rules. Most of the students had noticed that some classmates were less compliant with the restrictions and were critical of how peers met in larger groups and even threw parties. However, a few of the informants admitted that they occasionally bent the rules and met with more friends than recommended. They also said that some friends made excuses for being together by arguing that if any of them had the virus, they would have spread it to the rest already.

### Missing social interactions

How the participants reported their well-being depended on the magnitude of restrictions. The informants living with fewer regulations described that after the lockdown in spring 2020 was ended, much of their everyday life went on as normal, although their leisure time was more boring. The adolescents living with stronger social regulations and less physical attendance at school, said straight out that their quality of life was low and that they felt lonely, bitter, sad and depressed. Most of the participants stated that the number one cause of the decreased quality of life was that they were restricted from being with their friends and their extended family. When the schools closed down in spring 2020, they realized the importance of school, not only as an arena for learning but also for socializing:

> *“Normally, you don’t understand how much you need it (the school), how important it is, until you kind of loose it. Shortly after everything closed down and I couldn’t go to school, I just wanted to go there and meet my friends*.*” (Boy, group 2)*

A recurring topic was how the social restrictions resulted in difficult and awkward situations. Several described the unpleasant experience of not allowing someone to come over because the limit of guests already was reached. The almost impossible task of having to choose between friends was described by one boy:

> *“What I think is really hard is that I have to choose who to be with. And then someone says: “Oh, why did you choose those guys?” So you have to decide who you like the most, but you don’t want to do that either*.*” (Boy, group 4)*

While a few of the participants said that the social restrictions had made them more creative in their use of social media platforms, they all agreed that social media could not make up for human contact. One girl had observed that it is even harder to make contact online with someone you have not seen for a long time.

### Loss of opportunities

The participants were concerned that the constant shifting between action levels at school and the lack of continuity would negatively affect their learning outcome and future studies. Most of the adolescents said that online teaching was uninspiring. Some said that the teacher did not see them and how lost they felt. Instead, they were given too many assignments and too little assistance. Several said that their grades had dropped significantly over the course of the year.

Some of the adolescents expressed a sense of stagnation and that every day was the same. Attending online classes and spending the entire day indoors in their bedroom made them feel sleepy and apathetic. Much of their misery seemed to derive from having nothing to look forward to—no parties, no concerts, and no vacations. At the time of the pandemic outbreak, these adolescents had started to experience the freedom of making their own decisions and becoming more independent, and then suddenly they were prohibited from engaging in everything that is fun. For those living under strong restrictions and without access to leisure time activities, the situation seemed particularly hard. One boy described staying at home doing nothing as totally meaningless. Others spoke about lost opportunities, and feeling that they were missing something potentially significant for their development:

> *“Often, I think that what if there was no corona, what would I have experienced? We were barely halfway through our first year in upper secondary when… I just feel that I don’t grow as a person, I’m stuck in the first year and then time goes by and suddenly upper secondary school will be over*.*” (Boy, group 4)*

## Discussion

The participating adolescents stood out as very informed about the protective measures and were highly obedient to the government’s regulations. They were well aware of the dos and don’ts and noticed when others failed to comply with the advice and directives. The participants’ descriptions of how the protective measures were translated into their everyday lives demonstrate that they took responsibility and accepted personal costs for collective benefits. However, it seems evident that the situation negatively affected their quality of life, particularly due to social restrictions.

### Official information sources were beacons in the ocean of information

In the early phase of the pandemic, adolescents reported that traditional media were their most frequently used source of pandemic-related information [2]. Several months later, this was still the case among the interviewed adolescents. They emphasized watching the government’s and health authorities’ regular press conferences broadcast on national TV. Such briefings are the public’s first opportunity to be informed about new regulations. The importance of the health authorities’ communication channels underscores how important it was for the adolescents to get immediate and trustworthy updates and instructions for how their following weeks should be organized with regards to school, leisure activities and social interactions. This illustrates the abnormality of the entire situation, that young people were sitting at home waiting for governmental approval to be with friends or go to school. Official websites, including school websites, were among the sources the adolescents actively searched for information. The National Institute of Public Health’s website was frequently mentioned as the go-to place for accurate information on national regulations. Before the pandemic, most adolescents were probably not aware of this institution, which demonstrates the position public health authorities have taken also among young people. Social media have become significant communication tools for governments, organizations, universities, and schools to distribute crucial information to the public during the pandemic [18]. The Norwegian health authorities’ attempt to reach young people through social media seemed to work and the adolescents were attentive to the official health authorities as the sender of these messages. Except for watching the news and accessing the aforementioned websites, the adolescents gave the overall impression of being receivers of information and directives and that the information flow was massive. Protective behavior like washing hands and keeping a distance had become a habit. However, navigating the stream of sometimes contradictory information provided by national and local authorities about the number of people allowed to meet, challenged their comprehension of how to act.

### Compliance with protective measures despite confusion and frustration

Being health-literate in the context of a pandemic does not solely imply being able to find and understand information about protective measures. It also means being able to grasp the reasons behind the recommendations and regulations and to reflect on the outcomes of various actions [19, 20]. The adolescents in this study actively deliberated on the consequences of following the regulations or not, but were also confused about the rationale for some of the local rules at school. Despite frustration and despair, there was little opposition to the basic protective measures. Citizen trust in government is generally high in Norway and also among youth [21]. During spring 2020, trust in the government’s handling of the pandemic increased even more [22] which resonates with the overall loyal reaction to the health authorities’ appeal to engage in collective efforts to reduce the spread of the virus in the pandemic’s first phase [23, 24]. In the present study, the adolescents still stood out as concerned, showing solidarity with people vulnerable to the disease and being willing to help out. This is not unique for Norwegian adolescents. Motivation to engage in preventive behaviors based on concern for others is also described elsewhere [3, 5].

### Quality of life degraded by missed opportunities and social deprivation

In our survey study, we found a striking decrease in quality of life compared to European norm data showing that the pandemic has most certainly negatively affected the adolescents’ well-being [2]. In the present interview study, the adolescents confirmed the previous findings by describing their quality of life as low while also elaborating on factors significant for the decrease. The most socially restricted adolescents gave the richest descriptions of how impaired their everyday life had become. Having nothing to look forward to and plan for was described as one major cause for the adolescents’ decrease in quality of life. Quality of life is defined by the World Health Organization as individuals’ perception of their position in life in the context of their culture and value systems in which they live and in relation to their goals, expectations, standards, and concerns [25]. Pursuit of intrinsic aspirations, such as personal growth and meaningful relationships, foster well-being [26]. However, the pandemic has set restrictions to peoples’ autonomy and their ability to satisfy their needs for meaningful experiences, to make plans and set goals for the near future. Previous research has found that purpose embedded in the meaning of life concept is central to adolescent well-being [27]. Engaging in preventive behavior to protect relatives and vulnerable groups was perceived as meaningful by the adolescents in our study, and perhaps the main cause for still loyally following the guidelines. However, after several months of limitations, the engagement from the first lockdown seemed to have been replaced by apathy and resignation among some of the adolescents. The rapid change from having an eventful and increasingly independent life to spending a lot more time at home caused boredom, also described by adolescents in other studies [28, 29]. One may argue that things will normalize when the pandemic is over. However, not knowing how long the pandemic and the restrictions will last may have increased the feeling of distress. Also, it is important to recognize the adolescents’ concerns with what is potentially lost when their access to school is restricted, both with regards to their academic and their personal development. The long-term effects of losing in-person schooling have yet to be investigated, but, according to the adolescents in our study, and in other studies [30, 31], not being at school has negatively affected their work habits, motivation, learning gains and social life.

In a qualitative study exploring adolescents’ perceptions of what matters most to their quality of life, being with friends stood out as the most important [32]. Unsurprisingly, missed opportunities for social interactions with friends were the most prominent topic in the interviews. This corresponds with findings from qualitative studies carried out in other countries in which adolescents reported that not being with friends was among their biggest challenges [28-30]. Adolescence is a period of increased need for peer interaction, and the negative effects of physical distancing and social deprivation may have profound consequences [33]. Although adolescents in Norway have experienced fewer restrictions compared to adolescents in many other countries, the participants in our study pointed out that the limitations set on their life made them lonely and miserable. A clear association has previously been found between loneliness and mental health problems in adolescents [7]. Similar findings were reported in a Norwegian study investigating the association between loneliness and symptoms of anxiety and depression among adolescents during the pandemic [34].

It is reasonable to argue that adolescents’ extensive use of different social media applications makes them more resilient to social restrictions compared to children, adults and elderly people. Directed digital interaction through social media, for example, comments, likes, status updates and chatting with friends, has been found to be associated with greater well-being and lower loneliness [35]. Thus, use of digital applications may mitigate the potentially harmful effects of physical distancing [36]. However, the adolescents in the current study found social media to be insufficient compensation for meeting friends in real life. Moreover, observing on social media platforms that peers were socializing and even partying despite restrictions, was experienced as frustrating. Previous research has shown that passive usage of social media sites may elicit social comparisons and envy and thus cause distress [37]. The pandemic adds a new dimension to this by giving the adolescents glimpses of a life they craved but could not have, unless they too decided to act irresponsibly and break the rules.

### Strengths and limitations

In this study, we did not include member checking, which may have reduced the study’s response validity. It is a limitation that our sample did not include immigrant youth living in the most deprived parts of the capital, which have the highest prevalence of detected cases of Covid-19. Thus, we have recommend future studies to elaborate on the experiences of these minority adolescents. Due to the ongoing pandemic, we had to organize digital focus groups. Having a conversation online about potentially sensitive subjects may have been challenging thus affecting the participants’ responses. One way of minimizing potential discomfort and increasing a feeling of safety was to arrange for friends to be part of the same focus group. A strength of our study is that we recruited participants from different parts of Norway living with different restrictions. Using the findings from the previous survey gave valuable directions for the interview guide and analysis in the process of conducting this qualitative study.

## Conclusion

The adolescents gave the overall impression of being health literate which corresponds with the results from our previous survey study. During the second phase of the Covid-19 pandemic, protective measures had become a part of their everyday life. They knew where to find reliable information, they were attentive to the sender of information, and they tried their best to follow the rules and advice to prevent transmission. The adolescents seemed to accept that, in a pandemic, population health needs should be prioritized over the individual’s needs. However, they were clear about the social restrictions’ negative consequences for their quality of life and related this to missed opportunities to engage in important life events, interrupted schooling and not being able to freely socialize with friends. This study adds to the limited qualitative research on adolescents’ pandemic-related experiences and gives important insight into the burdens experienced by adolescents during the pandemic as well as their capabilities and resources.

## Data Availability

Data cannot be shared publicly because it consists of text from focus group interviews that is all in Norwegian.

## Acknowledgements

We wish to thank the participating adolescents for their valuable contributions to this study.

